# Wealth Effects of the Decrease in Under-five Mortality in India, 2005-2021

**DOI:** 10.1101/2023.04.11.23288395

**Authors:** Aalok Ranjan Chaurasia

**Affiliations:** MLC Foundation and ‘Shyam’ Institute

## Abstract

The present paper analyses the contribution of different population sub-groups classified by the wealth index quantiles groups to the change in the risk of death in the first five years of life in India during 2005-06 through 2019-21. The analysis reveals that the poorest and the poorer population sub-groups have primarily contributed to the decrease in the risk of death in the first five years of life in India whereas the contribution of the contribution of the richest population sub-group has been negative during the period under reference. The analysis also reveals that the wealth effects of the decrease in under-five mortality in different states of the country are different and, in many states, the contribution of the richest population sub-group to the decrease in the risk of death in the first five years of life in the state has been negative because of the change in the proportionate share of currently married women of reproductive age across different population sub-groups and their fertility. The paper emphasises the need of reinvigorating the health care services delivery system of the country so as to prevent under five deaths which cannot be prevented through the low-cost appropriate medical technology.

## Introduction

Wealth effects of under-five mortality are well-known (Biradar et al, 2018; Chao et al, 2018; Lartey et al, 2016). The probability of death in the first five years of life is the highest in the poorest but the lowest in the richest sub-group of the population. The decrease probability of death in the first five years of life is also known to be sensitive to the level of the probability of death in the first five years of life. The decrease is more rapid when the probability of death is high compared to the decrease when the probability of death is low. The reason behind the sensitiveness in the decrease with the level of the probability of death in the first five years of life is attributed to the difference between the structure of the causes of under-five deaths when the probability of death is high as compared to when the probability of death is low. The causes of under-five deaths may broadly be categorised into two groups – causes of death that can be prevented through the low-cost, appropriate medical technology like immunisation and oral rehydration therapy and causes of under-five deaths which cannot be prevented through the low-cost appropriate medical technology. Following the terminology first used by Bourgeois-Pichat (1952), the first group of the causes of under-five deaths may be termed as the ‘soft’ rock of under-five mortality while the second group of under-five deaths may be termed as the ‘hard’ rock of under-five deaths. The epidemiological transition theory suggests that as the under-five mortality decreases, the proportion of under-five deaths which cannot be prevented through the low-cost appropriate medical technology decreases while the proportion of under-five deaths that cannot be prevented through the low-cost appropriate medical technology increases. This essentially means that with the decrease in the probability of death in the first five years of life, it becomes increasing difficult to decrease the probability of death in the first five years of life further. The decrease in the probability of death in the first five years of life is, therefore, slow when the prevailing probability of death in the first five years of life is low as compared to when the probability of death in the first five years of life is high. Since the probability of death in the first five years of life is inversely related to the standard of living, the foregoing discussions imply that the relative contribution of the decrease in the probability of death in the first five-years of life in the poorest sub-group of the population will be more than the decrease in the probability of death in the first five years of life in the richest population sub-group. This conclusion ignores the fact that the decrease in the probability of death in the first five years of life is not evenly distributed in terms of the difficulty in decreasing the probability of death further on the scale of the probability of death in the first five years of life. For example, a drop of 10 points from an initial level of the probability of death in the first five years of life of 0.080 can be achieved much more easily than an equivalent drop from an initial level of the probability of death in the first five years of life of 0.035. From the analytical perspective, it is, therefore, important that the uneven distribution of the difficulty in further decreasing the probability of death in the first five years of life on the scale of the probability of death in the first five years of life is taken into account while analysing the relative contribution of the decrease in the probability of death in the first five years of life in different population sub-groups.

The contribution of the decrease in the probability of death in the first five years of life in different population sub-groups to the decrease in the probability of death in the first five years of life in the population is also influenced by the change in the distribution of live births across different population sub-groups. The distribution of live births across population sub-groups, in turn, is influenced by the change in the proportionate distribution of currently married women of reproductive age across different population sub-groups and their fertility relative to the average fertility of all currently married women of reproductive age. Like the probability of death in the first five years of life, the fertility of currently married women of reproductive age is also inversely related to the standard of living – poorest women have the highest fertility while the richest women have the lowest fertility, in general. On the other hand, with the process of social and economic development, the proportion of currently married women of reproductive age in the poorest and the poorer population sub-groups decreases while the proportion of currently married women of reproductive age in the richest and the richer population sub-group increases. This shift the in the proportionate distribution of the currently married women of reproductive age by the standard of living or the level of income has implications for the distribution of live births across different population sub-groups and hence to the contribution of the decrease in the probability of death in the first five years of life in different sub-groups of the population to the decrease in the probability of death in the first five years of life in the whole population.

In this paper, we analyse the wealth effects of the decrease in the probability of death in the first five years of life in India during the period 2005-06 through 2019-21 following a decomposition approach. The analysis is based on the probability of death in the first five years of life (_*5*_*q*_*0*_) and a transformation of _*5*_*q*_*0*_ which ensures a more even distribution of the difficulty in further decreasing _*5*_*q*_*0*_ with the decrease in _*5*_*q*_*0*_. The approach that we have used estimates the contribution of the change in _*5*_*q*_*0*_ in five population sub-groups in terms of the standard of living – poorest, poorer, middle, richer and richest – to the change in _*5*_*q*_*0*_ in the population between 2005-06 and 2019-21. We find that when the uneven distribution of the difficulty in further decreasing _*5*_*q*_*0*_ with the decrease in _*5*_*q*_*0*_ is taken into account, the contribution of the change in _*5*_*q*_*0*_ in different population sub-groups to the change is _*5*_*q*_*0*_ in the population is different when the uneven distribution of the difficulty in further decreasing _*5*_*q*_*0*_ with the decrease in _*5*_*q*_*0*_ is not taken into consideration.

The rest of the paper is organised into five sections. The next section of the paper describes the data used in the analysis. The analysis is based on the data available from the third round (2005-06) and the fifth round (2019-21) of the National Family Health Survey. The third section of the paper describes the decomposition methodology used in analysing the contribution of the change in _*5*_*q*_*0*_ in different population sub-groups to the change in _*5*_*q*_*0*_ in the population between 2005-06 and 2019-21. Findings of the analysis are presented in section four of the paper for India and for its selected states. The key findings of the analysis are discussed from the policy and the programme perspective in the fifth and the last section of the paper.

## Data

The analysis is based on the third and the fifth rounds of the National Family Health Survey (NFHS) in India. The third (2005-06) round of the survey provides full birth histories of a nationally representative sample of 124385 currently married women of reproductive age (15-49 years). The sample covered 99 per cent of the population of the country living in 29 states and Union Territories (Government of India, 2007). The fifth (2019-21) round of NFHS, on the other hand, provides full birth histories of a nationally representative sample of 724115 currently married women of reproductive age and covers the entire country (Government of India, 2021). In both rounds of the survey, the wealth status of the women interviewed was assessed based on a composite wealth index score constructed from a set of household level assets, amenities, and facilities. Based on this composite wealth index score, all currently married women of reproductive age have been classified into one of the five wealth index quintiles groups – poorest, poorer, middle, richer and richest. Using the full birth history data of the currently married women of reproductive age, estimates of the probability of death in the first five years of life (U5MR) was prepared for the period five years preceding the survey were calculated for each of the five wealth index quintiles groups using the CMRJack software for the estimation of child mortality (Pederson and Liu, 2012). In addition, fertility of the currently married women of reproductive age (general marital fertility rate) in each of the five wealth index quintiles group has also been calculated based on the number of live births reported during the five years period preceding the survey. These estimates constituted the basic dataset for the present analysis.

## Methods

Let *u* denotes a measure of the risk of death in the first five years of life and *u*_*i*_ denotes the risk of death in the first five years of life in population sub-group *i, i*=1 (Poorest), 2 (Poorer), 3 (Middle), 4 (Richer), and 5 (Richest). Then, we can write,

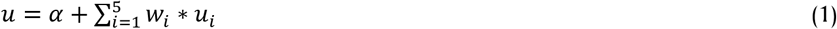

Here *w*_i_ is the proportion of live births in the population sub-group *i* to the total live births in the population so that.

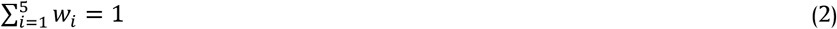

The value of *α* in equation (1) should ideally be equal to zero but, practically, there is always some difference between *u* and weighted sum of *u*_*i*_ because the standard of living of the currently married women of reproductive age is not the only factor that determines both _*5*_*q*_*0*_ and the fertility of the currently married women.

The above formulation suggests that the change in *u* between two points in time, *t*^*2*^ and *t*^*1*^, can be decomposed as

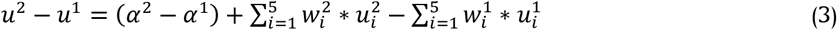

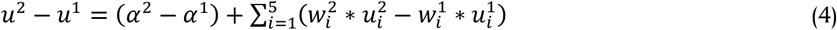

We can write,

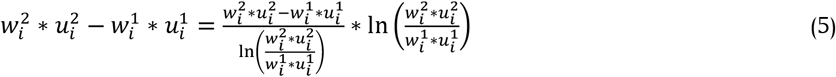

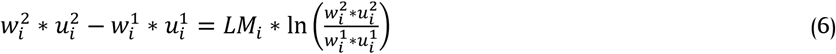

Here

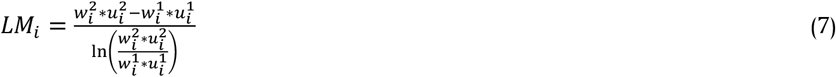

is the logarithmic mean (Carlson, 1974).

Now

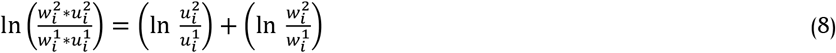

If *L* denotes the number of live births, *M* denotes the number of currently married women of reproductive age and *G* denotes the general marital fertility rate in the population while *L*_*i*_, *M*_*i*_ and *G*_*i*_ denote, respectively, the number of live births, number of currently married women of reproductive age and general marital fertility rate in the population sub-group *i*, then, we can write

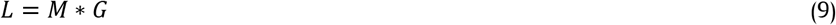

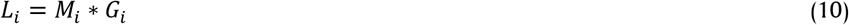

Now

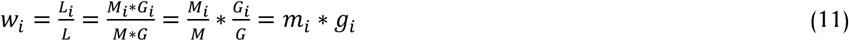

Here *m*_*i*_ is the proportion of currently married women of reproductive age in the population sub-group *i* and *g*_*i*_ is the ratio of the general marital fertility rate in the population sub-group *i* to the general fertility rate in the population. Equation (8) can now be written as

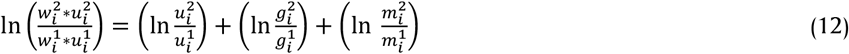

Substituting, we get

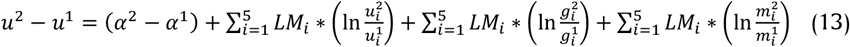

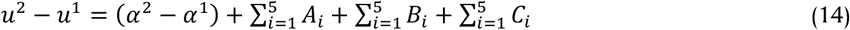

or

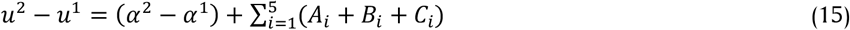

where

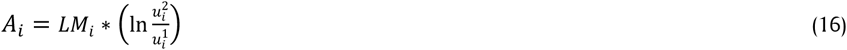

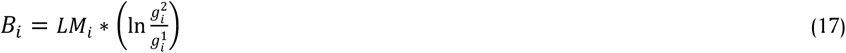

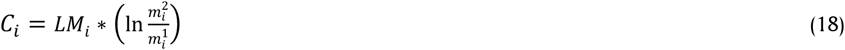

Equation (15) suggests that the change in the risk of death in the first five years of life (*u*) between two points in time can be decomposed into four components - one attributed to the change in the risk of death in the first five years of life in different population sub-groups (*A*_*i*_), second attributed to the change in the general marital fertility rate in different population sub-groups (*B*_*i*_), third attributed to the change in the proportionate share of currently married women of reproductive age in different population sub-groups (*C*_*i*_), and fourth attributed to the change in other factors that influence the risk of death in the first five years of life that is not accounted by the standard of living of currently married women of reproductive age. Equation (15) also suggests that the contribution of a population sub-group to the change in the risk of death in the first five years of life in the population is not determined by the change in the risk of death in the first five years of life in the population sub-group alone but also by the change in the proportionate share of currently married women of reproductive age and the change in the fertility of currently married women of reproductive age.

Application of equation (15) requires a measure of the risk of death in the first five years of life. The most commonly used measure is _*5*_*q*_*0*_. However, as discussed earlier, _*5*_*q*_*0*_ is sensitive to uneven distribution of the difficulty in further decrease in _*5*_*q*_*0*_ with the decrease in _*5*_*q*_*0*_. It is more appropriate to use a measure that ensures a more even distribution of the difficulty in further decrease in the risk of death in the first five years of life with the decrease in the risk of death in the first five years of life. Following Mosteller and Tukey (1977), such a measure of the risk of death in the first five years of life can be constructed by an appropriate transformation of _*5*_*q*_*0*_. Thus, a transformation of _*5*_*q*_*0*_, to be termed as the under-five mortality index (_*5*_*i*_*0*_) may be defined as:

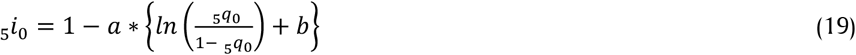

The index _*5*_*i*_*0*_ distributes the degree of difficulty in achieving further decrease in _*5*_*q*_*0*_ with the decrease in _*5*_*q*_*0*_ more uniformly. Here, the parameters *a* and *b* may be determined in such a manner that _*5*_*i*_*0*_=0 corresponding to the minimum possible value of _*5*_*q*_*0*_ and _*5*_*i*_*0*_=1 corresponding to the maximum possible value of _*5*_*q*_*0*_. We assume that _*5*_*q*_*0*_ ranges from a minimum possible value of 0.003 to the maximum possible value of 0.025. These limits of _*5*_*q*_*0*_ give *a*=-0.20166 and *b*=0.84730. If the minimum and maximum possible values of _*5*_*q*_*0*_ are changed, the values of the parameters *a* and *b* in equation (19) will change.

We have measured the standard of living of the currently married women of reproductive age in terms of the wealth index which is based on household assets and utility services. The details of the construction of the wealth index are given elsewhere and not repeated here (Rutstein and Johnson, 2004). Based on the distribution of the wealth index, currently married women of reproductive age have been classified into five sub-groups – poorest (women having wealth index lower than the first quintiles); poorer (women having wealth index between first and second quintiles); middle (women having wealth index between second and third quintiles); richer (women having wealth index between third and fourth wealth quintiles); and richest (women having wealth index higher than the fourth quintiles).

## Results

Table 1 gives estimates of _*5*_*q*_*0, 5*_*i*_*0*_, proportionate distribution of currently married women of reproductive ages (*m*_*i*_) and the fertility of currently married women of reproductive age (*GMFRI*) in different population sub-groups as revealed through the third round (2005-06) and the fifth round (2019-21) of the National Family Health Survey. The table also gives estimates of Theil’s inequality index across the five population sub-groups in _*5*_*q*_*0, 5*_*i*_*0*_, *m*_*i*_ and *GMFR*. The inequality in _*5*_*q*_*0*_ across the five population sub-groups is lower in 2019-21 compared to that in 2005-06 which suggests that the decrease in _*5*_*q*_*0*_ has been comparatively faster in the poorest and the poorer population sub-groups compared to that in richer and the richest population sub-groups. However, when the uneven distribution of the difficulty in decreasing _*5*_*q*_*0*_ further with the decrease in _*5*_*q*_*0*_ is taken into consideration, the inequality across the five sub-groups of the population, the inequality in _*5*_*i*_*0*_ shows an increase in 2019-21 compared to that in 2005-06. This means that the decrease in the inequality in _*5*_*q*_*0*_ across different population sub-groups between 2005-06 and 2019-21 is a reflection of the uneven distribution of the difficulty in further decreasing _*5*_*q*_*0*_ with the decrease in _*5*_*q*_*0*_. When the effect of the uneven distribution of the difficulty in further decreasing _*5*_*q*_*0*_ with the decrease in _*5*_*q*_*0*_ is controlled, the decrease in the risk of death in the first five years of life across different population sub-groups in India appears to have diverged. Tables 1 also suggest that the inequality across the five population sub-groups has also decreased in the proportionate distribution of currently married women of reproductive age and in fertility of these women.

**Table 1:** Estimates of _*5*_*q*_*0, 5*_*i*_*0*_, GMFR and proportionate distribution of currently married women of reproductive age by wealth quintiles in India, 2005-06 and 2019-21.

| Wealth quintiles | $sq_0$ | $si_0$ | GMFR | Proportion of currently married women of reproductive age |
| --- | --- | --- | --- | --- |
| 2005-06 |  |  |  |  |
| Poorest | 0.1005 | 0.7289 | 0.1726 | 0.2297 |
| Poorer | 0.0896 | 0.7033 | 0.1382 | 0.2221 |
| Middle | 0.0719 | 0.6550 | 0.1160 | 0.2032 |
| Richer | 0.0512 | 0.5821 | 0.1005 | 0.1862 |
| Richest | 0.0338 | 0.4947 | 0.0750 | 0.1589 |
| All | 0.0743 | 0.6622 | 0.1245 | 1.0000 |
| Theil's inequality index | 0.0656 | 0.0093 | 0.0381 | 0.0083 |
| 2019-21 |  |  |  |  |
| Poorest | 0.0581 | 0.6090 | 0.1131 | 0.2312 |
| Poorer | 0.0462 | 0.5605 | 0.0904 | 0.2174 |
| Middle | 0.0381 | 0.5200 | 0.0785 | 0.2031 |
| Richer | 0.0301 | 0.4705 | 0.0733 | 0.1879 |
| Richest | 0.0204 | 0.3897 | 0.0663 | 0.1605 |
| All | 0.0408 | 0.5342 | 0.0862 | 1.0000 |
| Theil's inequality index | 0.0580 | 0.0112 | 0.0182 | 0.0076 |

Source: Author

Table 1 also suggests that the general marital fertility rate (*GMFR*) has also decreased in all the population sub-groups, and the decrease has been the fastest in the poorest sub-group of the population but the slowest in the richest population sub-group. Like _*5*_*q*_*0*_, the difficulty in further decreasing *GMFR* with the decrease in *GMFR* is also unevenly distributed with the level of *GMFR*. In the poorest and the poorer sub-groups of the population, *GMFR* decreased by around 34 per cent between 2005-06 and 2019-21 whereas, in the richest population sub-group, *GMFR* decreased by only around 12 per cent during this period.

The proportionate share of the currently married women of reproductive age in the five population sub-groups has also changed between 2005-06 and 2019-21. The proportionate share of the poorest currently married women of reproductive age has increased but that of the poorer currently married women of reproductive age has decreased. There has been virtually no change in the proportionate share of currently married women of reproductive age in the middle wealth index quintiles group, but this share has increased in both richer and the richest population sub-groups between 2005-06 and 2019-21. This change in the proportionate share of the currently married women of reproductive age in different population sub-groups, along with the change in the fertility of currently married women of reproductive age, has resulted in a change in the proportionate share of live births in different population sub-groups and influences the contribution of the change in the risk of death in first five years of life in different population sub-groups to the change in the risk of death in the population.

The wealth effects of the change in the risk of death in the first five years of life are presented in table 2 when risk of death in the first five years of life is measured in terms of _*5*_*q*_*0*_. Almost 40 per cent of the decrease in _*5*_*q*_*0*_ in India between 2005-06 and 2019-21 the country is accounted by the decrease in _*5*_*q*_*0*_ in the poorest population sub-group whereas the poorer population sub-group has accounted for a decrease of 32 per cent. This means that the decrease in _*5*_*q*_*0*_ in the poor population has accounted for almost 72 per cent of the decrease in _*5*_*q*_*0*_ in the country between 2005-06 and 2019-21. By contrast, the decrease in _*5*_*q*_*0*_ in the richest population sub-group accounted for only around 2 per cent of the decrease in _*5*_*q*_*0*_ whereas the decrease in _*5*_*q*_*0*_ in the richer population sub-group has accounted for only around 8 per cent of the decrease in _*5*_*q*_*0*_ in the country between 2005-06 and 2019-21. In other words, the decrease in _*5*_*q*_*0*_ in the rich population of the country accounted for only about 10 per cent of the decrease in _*5*_*q*_*0*_ in the country between 2005-06 and 2019-21. Finally, the middle-income population sub-group has accounted for about 18 per cent of the decrease in _*5*_*q*_*0*_ in the country.

**Table 2:** Wealth effects of the change in _*5*_*q*_*0*_ in India, 2005-06 through 2019-21.

| Wealth quintiles | Contribution of the change in $U5MR$ in the wealth quintiles attributed to the change in | | | | |
| --- | --- | --- | --- | --- | --- |
| | $sq_0$ | $GMFR$ | Proportionate share of women | Total contribution | |
| Poorest | -0.0132 | -0.0013 | 0.0002 | -0.0144 | 39.8 |
| Poorer | -0.0103 | -0.0009 | -0.0003 | -0.0115 | 32.0 |
| Middle | -0.0063 | -0.0002 | 0.0000 | -0.0066 | 18.2 |
| Richer | -0.0033 | 0.0003 | 0.0001 | -0.0029 | 8.0 |
| Richest | -0.0014 | 0.0007 | 0.0000 | -0.0007 | 2.0 |
| Total | -0.0346 | -0.0014 | -0.0001 | -0.0361 | 100.0 |
| Contribution of change in other factors |  |  |  | 0.0026 |  |
| Total change in $sq_0$ in the population | | | | -0.0335 | |

Source: Author

Table 2 also shows how the change in _*5*_*q*_*0*_, *GMFR* and the proportionate share of currently married women of reproductive age in a population sub-group determine the contribution of that population sub-group to the change in _*5*_*q*_*0*_ in the population. The _*5*_*q*_*0*_ decrease in all population sub-groups, although the magnitude of the decrease has been different. In the poorest and in the middle-income population sub-groups, *GMFR* decreased between 2005-06 and 2019-21 but the proportionate share of the currently married reproductive age women increased so that the contribution of the change in _*5*_*q*_*0*_ in the two population sub-group has been less than that determined by the decrease in _*5*_*q*_*0*_ and the decrease in *GMFR*. In the richer and the richest population sub-groups, on the other hand, both *GMFR* and the proportionate share of currently married women of reproductive age increased, instead decreased, between 2005-06 and 2019-21 so that the contribution of these population sub-groups to the decrease in _*5*_*q*_*0*_ in the country has been less than that determined by the decrease in _*5*_*q*_*0*_. This leaves only the poorer population sub-group in which both *GMFR* and the proportionate share of currently married women of reproductive age decreased so that the contribution of this population sub-group to the decrease in _*5*_*q*_*0*_ in the country has been substantially higher than that determined by the decrease in _*5*_*q*_*0*_ in this sub-group of the population.

When _*5*_*i*_*0*_ is used as the measure of the risk of death in the first five years of life which ensures more equal distribution of the difficulty in further reduction in the risk of death with the decrease in the risk of death in the first five years of life, the decrease in _*5*_*i*_*0*_ in the poorest and the poorer population sub-groups accounted for around 70 per cent of the decrease in _*5*_*i*_*0*_ in the country whereas the decrease in _*5*_*i*_*0*_ in the richest and the richer population sub-groups accounted for a decrease of less than 9 per cent of the decrease in _*5*_*i*_*0*_ in the country (Table 3). Perhaps the most important observation of table 3 is that the contribution of the decrease in _*5*_*i*_*0*_ in the richest population sub-group to the decrease in _*5*_*i*_*0*_ in the country has negative, albeit marginal. This means that the decrease in _*5*_*i*_*0*_ in the richest population sub-group has actually contributed to increasing, not decreasing, _*5*_*i*_*0*_ in the country. The reason is that the contribution of the decrease in _*5*_*i*_*0*_ in the richest population sub-group has not been substantial enough to compensate for the contribution of the increase in *GMFR* and the proportionate share of currently married women of reproductive age.

**Table 3:** Wealth effects of the change in _*5*_*i*_*0*_ in India, 2005-06 through 2019-21.

| Wealth quintiles | Contribution of the change in $UMI$ in the wealth quintiles attributed to the change in | | | | |
| --- | --- | --- | --- | --- | --- |
| | $s_{i0}$ | $GMFR$ | Proportionate share of women | Total contribution | |
| Poorest | -0.0373 | -0.0113 | 0.0014 | -0.0472 | 35.8 |
| Poorer | -0.0339 | -0.0084 | -0.0032 | -0.0455 | 34.5 |
| Middle | -0.0253 | -0.0024 | -0.0001 | -0.0278 | 21.1 |
| Richer | -0.0173 | 0.0043 | 0.0008 | -0.0122 | 9.3 |
| Richest | -0.0114 | 0.0117 | 0.0005 | 0.0008 | -0.6 |
| Total | -0.1252 | -0.0061 | -0.0007 | -0.1319 | 100.0 |
| Contribution of change in other factors |  |  |  | 0.0039 |  |
| Total change in $s_{i0}$ in the population | | | | -0.1280 | |

Source: Author

It may also be seen from tables 2 and 3 that the marginal contribution of the decrease in _*5*_*q*_*0*_ and the negative contribution of the decrease in _*5*_*i*_*0*_ in the richest population sub-group to the decrease in _*5*_*q*_*0*_ or _*5*_*i*_*0*_ in the country may be attributed to the change in the proportionate distribution of currently married women of reproductive age and their fertility. The proportion of currently married women of reproductive age in the richest population sub-group increased from less than 15.9 per cent in 2005-06 to more than 16 per cent in 2019-21. On the other hand, although *GMFR* in the richest population sub-group decreased from 0.074 in 2005-06 to 0.066 in 2019-21, yet the *GMFR* in this population sub-group was only about 60 per cent of the *GMFR* of the country in 2005-06 but almost 77 per cent of the *GMFR* of the country in 2019-21 which means that the decrease in *GMFR* in this population sub-group has been slower than the decrease in *GMFR* in the country as a whole during this period. As a result of these changes in the proportionate share of the currently married women of reproductive age and their fertility, the proportion of live births in the richest population sub-group increased from less than 10 per cent to more than 12 per cent between 2005-06 and 2019-21 so that, despite the decrease in _*5*_*q*_*0*_ in this population sub-group, the contribution of the decrease in _*5*_*q*_*0*_ in this population sub-group to the decrease in _*5*_*q*_*0*_ in the country has been marginal. When _*5*_*i*_*0*_ is used as the measure of the risk of death in the first five years of life, the negative contribution of the change in the proportionate share of currently married women of reproductive age and their fertility in the richest population sub-group has been more than the positive contribution of the decrease in _*5*_*i*_*0*_ so that the contribution of the decrease in the risk of death in the first five years of life in the richest population sub-group to the decrease in _*5*_*i*_*0*_ in the country has been negative, not positive.

Table 4 presents the contribution of the change in _*5*_*q*_*0*_ in different population sub-groups to the change in _*5*_*q*_*0*_ in selected states of the country while table 5 presents the contribution of the change in _*5*_*i*_*0*_ in different population sub-groups to the change in _*5*_*i*_*0*_. Although, both _*5*_*q*_*0*_ and _*5*_*i*_*0*_ decreased in all population sub-groups in all the states included in the analysis, yet the contribution of different population sub-groups to the change in _*5*_*q*_*0*_ and _*5*_*i*_*0*_ in different states has been different. In Chhattisgarh and Jharkhand, for example, the change in _*5*_*q*_*0*_ in the poorest population sub-group accounted for more than 70 per cent of the change in _*5*_*q*_*0*_ in the state whereas this proportion was less than 25 per cent in Andhra Pradesh, Bihar, Gujarat, and Karnataka. On the other hand, in all states except Maharashtra and Madhya Pradesh, the contribution of the richest population sub-group to the change in _*5*_*q*_*0*_ in the state has been less than 10 per cent of the total change. Maharashtra is the only state where almost 19 per cent of the change in _*5*_*q*_*0*_ in the state between 2005-06 and 2019-21 is attributed to the richest population sub-group whereas Madhya Pradesh is the only state where the contribution of the richest population sub-group to the decrease in _*5*_*q*_*0*_ in the state has been negative meaning that the richest population sub-group in Madhya Pradesh contributed to the increase, instead decrease, in _*5*_*q*_*0*_ in the state between 2005-06 and 2019-21. The reason is that the contribution of the decrease in _*5*_*q*_*0*_ in the richest population sub-group of the state was not substantial enough to compensate for the negative contribution resulting from the increase in *GMFR* and the increase in the proportionate share of the currently married women of reproductive age in the richest population sub-group.

**Table 4:**
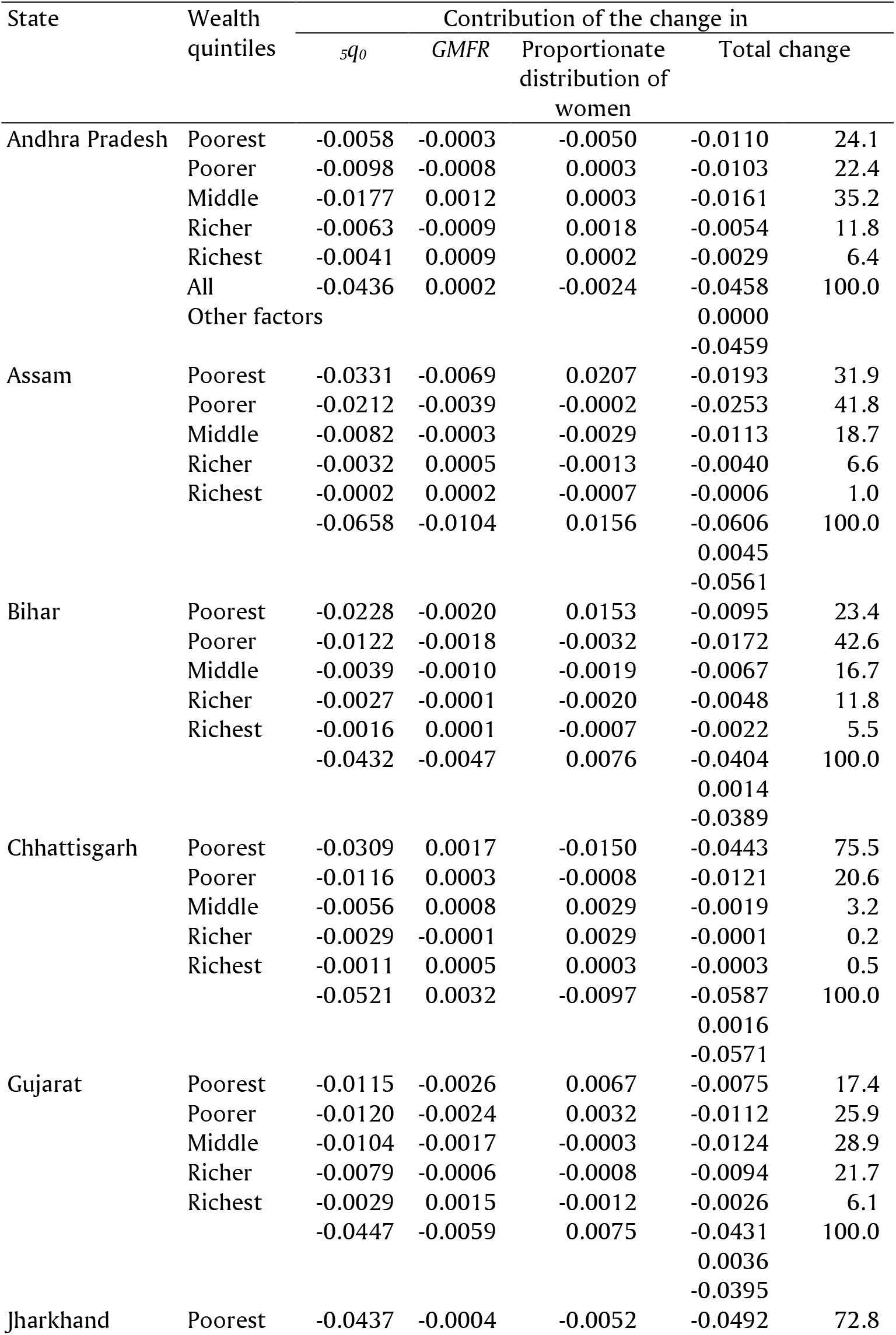
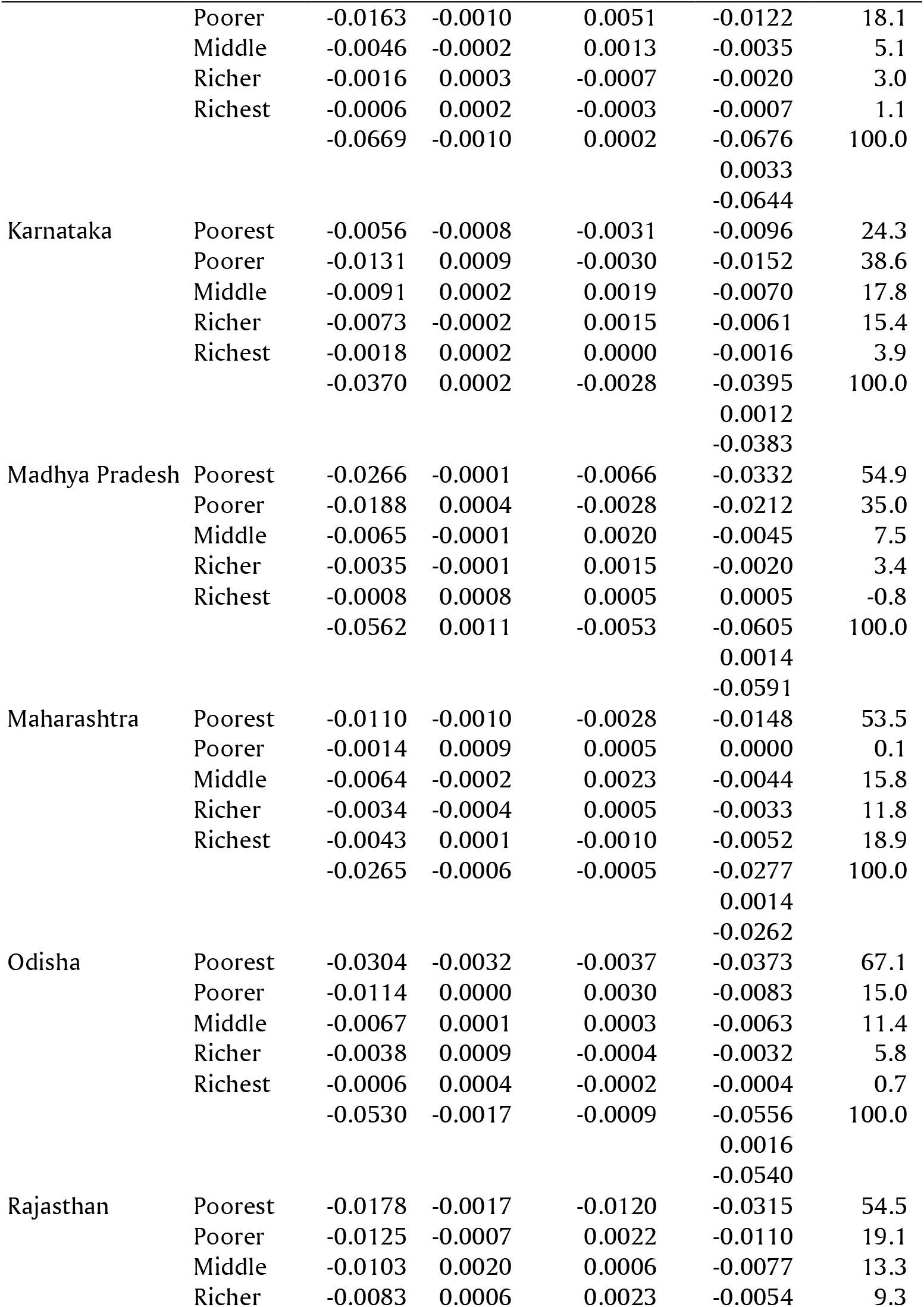
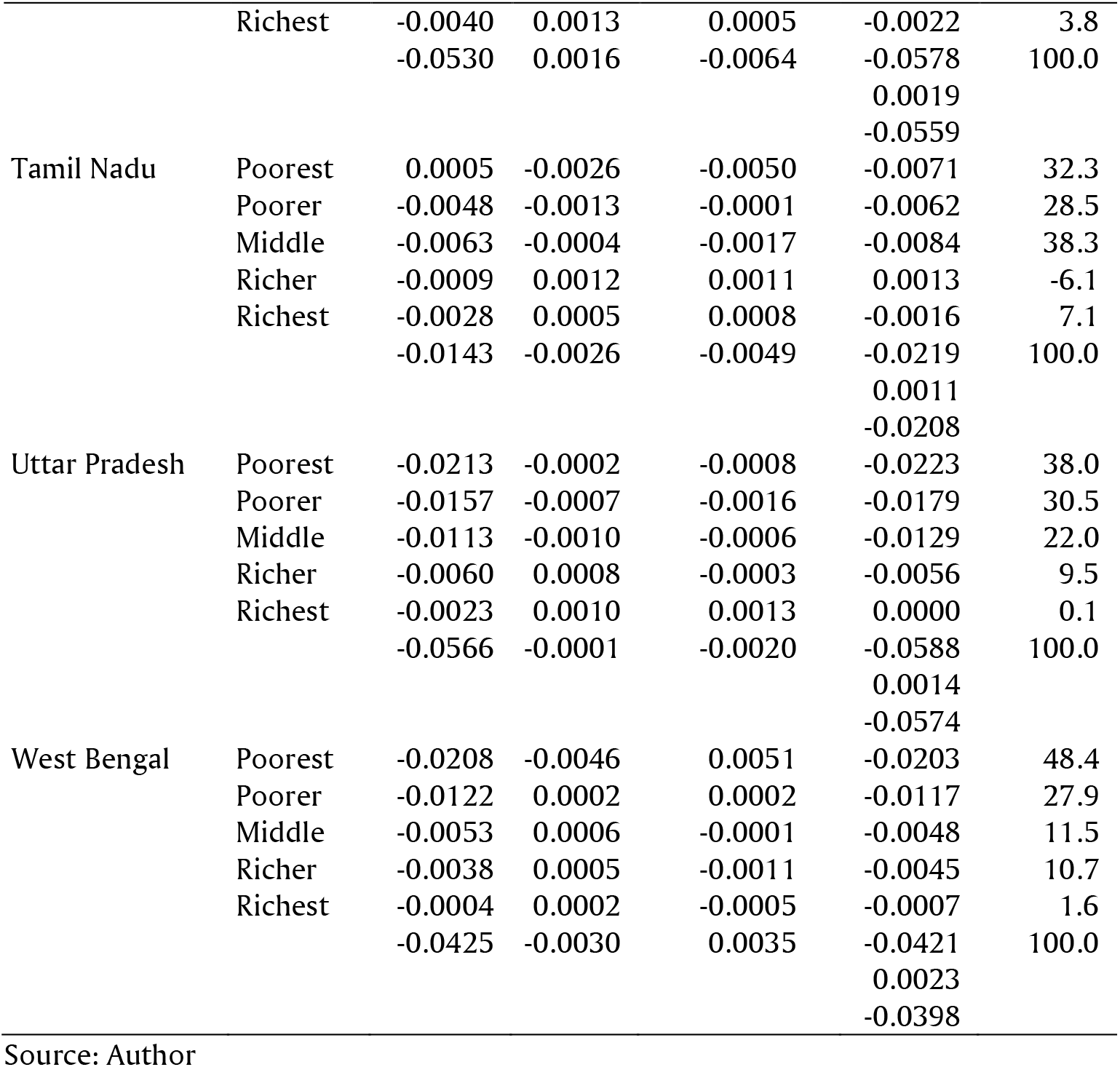
Wealth effects of the change in _*5*_*q*_*0*_ in selected states of India between 2005-06 and 2019-21.

**Table 5:**
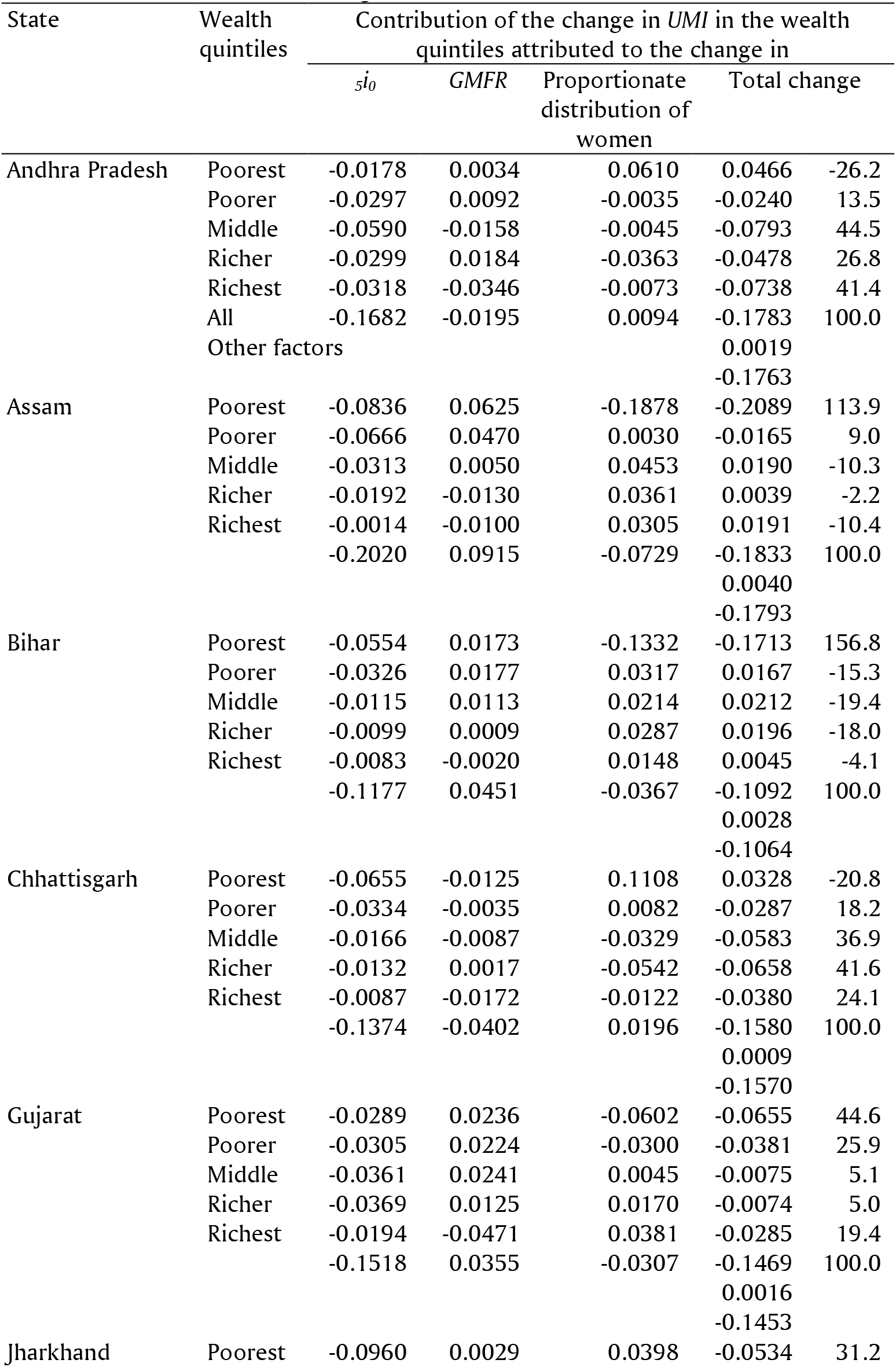
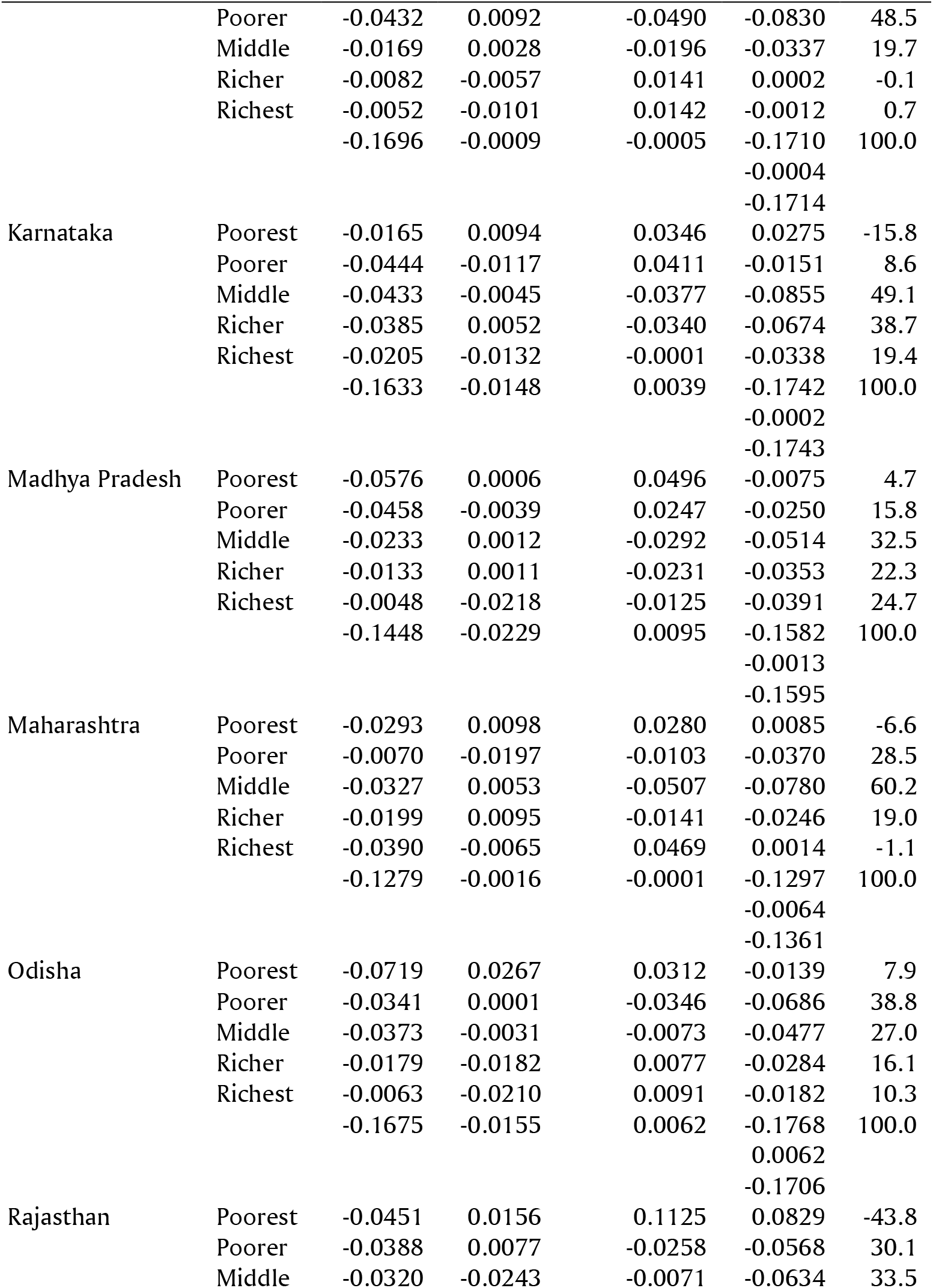
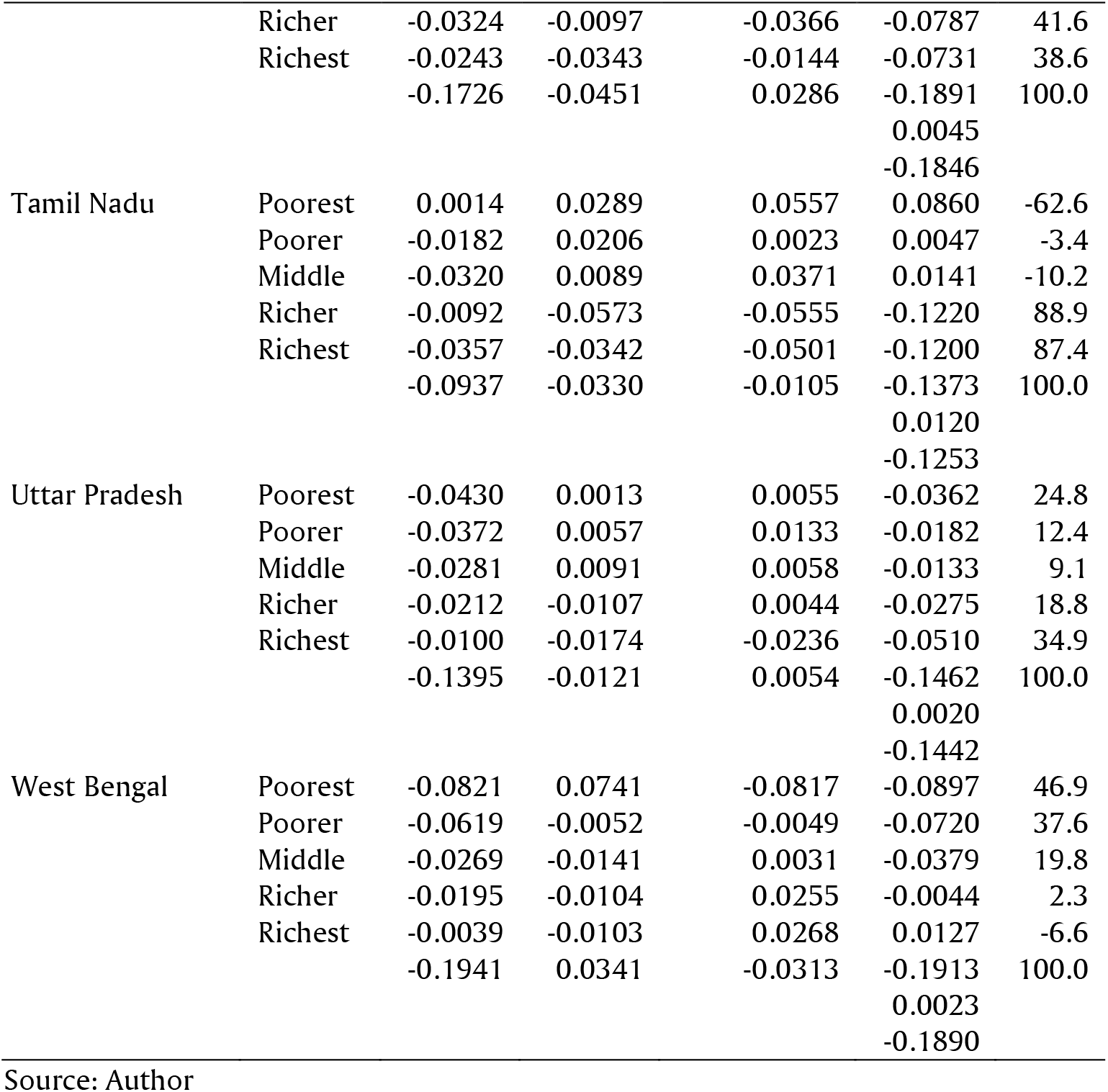
Wealth effects of the change in _*5*_*i*_*0*_ in selected states of India, 2005-21.

Table 4 also suggests that the pattern of change in _*5*_*q*_*0*_, *GMFR* and in the proportionate share of the currently married women of reproductive age in different population sub-groups has been different in different states. In all states, _*5*_*q*_*0*_ decreased in all population sub-groups between 2006-06 and 2019-21 with the only exception of Tamil Nadu where _*5*_*q*_*0*_ increased in the poorest population sub-group during this period. This has, however, not been the case with the change in *GMFR* and the change in the proportionate share of currently married women of reproductive age. The *GMFR* increased in the richest population sub-group in all states whereas it decreased in the poorest population sub-group in all states except Chhattisgarh. In other population sub-groups also, there has been no consistent pattern in the change in *GMFR* across the states of the country. The same has been the case with the proportionate share of currently married women of reproductive age. In Andhra Pradesh, the proportionate share of currently married women of reproductive age decreased in the poorest population sub-group only but increased in all other population sub-groups. In Assam, Bihar, and Rajasthan, on the other hand, the proportionate share of the currently married reproductive age women increased in the poorest population sub-group but decreased in all other population sub-groups.

The scenario is different when _*5*_*i*_*0*_ is used to measure of the risk of death in the first five years of life in place of _*5*_*q*_*0*_ (Table 5). In Andhra Pradesh, Chhattisgarh, Karnataka, Maharashtra, Rajasthan, and Tamil Nadu, for example, the contribution of the poorest population sub-group to the decrease in _*5*_*i*_*0*_ in the state has been negative which means that the poorest population sub-group in these states has actually contributed to increasing, instead decreasing the _*5*_*i*_*0*_. On the other hand, the decrease in _*5*_*i*_*0*_ between 2005-06 and 2019-21 in Bihar has been entirely due to the contribution of the poorest population sub-group as all the other population sub-groups in the state has contributed towards increasing rather than decreasing _*5*_*i*_*0*_ during the period under reference. By contrast, the entire decrease in _*5*_*i*_*0*_ in Tamil Nadu is attributed to the contribution of the richer and the richest population sub-groups as the contribution of the remaining three population sub-groups to the decrease in _*5*_*i*_*0*_ in the state has been negative. Table 5 also confirms that the contribution of different population sub-groups to the decrease in _*5*_*i*_*0*_ in different states has been different.

## Discussion and Conclusions

The present analysis reveals that the decrease in the risk of death in the first five years of life during the period 2005-06 through 2019-21 is primarily attributed to the contribution of the poorest and the poorer population sub-groups whereas the contribution of the richer and the richest population sub-groups to the decrease in the risk of death in the first five years of life has been marginal. There has been a decrease in the risk of death in the first five years of life in the richer and the richest population sub-groups measured in terms of either _*5*_*q*_*0*_ or _*5*_*i*_*0*_ but most of the contribution of this this decrease in the risk of death in the first five years of life is compensated by the negative contribution of the change in the proportionate share of the currently married women of reproductive age and their fertility in the richer and the richest population sub-groups. The analysis also reveals that when the difficulty in reducing the risk of death in the first five years of life with the decrease in the risk of death is distributed more equally, the contribution of the decrease in the risk of death in the first five years of life in the richest sub-group of the population to the decrease in the risk of death in the first five years of life in the country turns negative despite the decrease in the risk of death in the first five years of life because of the compositional changes in the proportionate share of the currently married women of reproductive age and changes in their fertility. A similar situation prevails in many states of the country where the decrease in the risk of death in the first five years of life in the richest population sub-group has not been large enough to compensate for the negative contribution resulting from the change in the proportionate share of the currently married women of reproductive age and the change in the fertility of these women. The risk of death in the first five years of life in the richest population sub-group was quite low even in 2005-06 which implies that most of the child deaths in this population sub-group were due to causes of death which cannot be prevented through the low-cost appropriate medical technology like immunisation and oral rehydration therapy. Prevention of these deaths require availability of and access to advanced medical technology. However, very little is known at present about the causes of under-five deaths in different population sub-groups in the country.

India has traditionally followed the low-cost appropriate medical technology approach to prevent avoidable under-five deaths and reduce the risk of death in the first five years of life. This approach has been quite successful in preventing those under-five deaths which can be prevented through such interventions as immunisation against vaccine preventable diseases and prevention of deaths due to dehydration in diarrhoea through simple oral rehydration therapy. With the success in the prevention under-five deaths through the application of the low-cost appropriate medical technology, the share of these under-five deaths is decreasing while the share of those under five death which cannot be prevented through the low-cost appropriate medical technology is increasing. The present analysis suggests that efforts to prevent under-five deaths in the country should go beyond the universalisation of the low-cost appropriate medical technology so as to prevent those under-five deaths which are beyond the ambit of the low-cost appropriate medical technology. The health care delivery system of the country needs to be reinvigorated in this context.

An important finding of the present analysis is that the wealth effects of the change in the risk of death in the first five years of life are not the same in different states of the country and, in different states of the country, the decrease in the risk of death in the first five years of life in different states has been confined to different population sub-groups classified on the basis of the wealth index. Bihar and Tamil Nadu are two extremes. In Bihar, the entire decrease in the risk of death in the first five years of life has been contributed by the poorest population sub-groups whereas, in Tamil Nadu, the entire decrease in the risk of death in the first five years of life has been confined to the richest sub-group of the population. It appears that there are state specific factors that influence the wealth effects of the change in in the risk of death in the first five years of life. Identification of these factors is important to ensure that the decrease in the risk of death in the first five years of life is distributed across different population sub sub-groups classified by the wealth index more uniformly. There is a need to examine the causes of under-five deaths in different population sub-groups in different states of the country.

The analysis also suggests that the uneven distribution of the difficulty in decreasing further the risk of death in the first five years of life with the decrease in the risk of death must be addressed before analysing the wealth effects of the change in the risk of death in the first five years of life. This requires alternate measures of the risk of death in the first five years of life than the conventional probability of death in the first five years of life. The wealth effects of the change in the risk of death in the first five years of life are radically different when the difficulty in reducing the risk of death in the first five years of life further with the decrease in the risk of death is not distributed more uniformly on the scale of the risk of death in the first five years of life. The present paper has used a transformation of the conventional probability of death in the first five years of life to distribute, more uniformly, the difficulty in further reducing the risk of death in the first five years of life with the decrease in the risk of death.

## Data Availability

Data can be made available upon request

